# Alterations in the Nasopharyngeal Microbiome Associated with SARS-CoV-2 Infection Status and Disease Severity

**DOI:** 10.1101/2022.06.13.22276358

**Authors:** Nick P.G. Gauthier, Kerstin Locher, Clayton MacDonald, Samuel D. Chorlton, Marthe Charles, Amee R. Manges

## Abstract

**Objectives:** The COVID-19 pandemic and ensuing public health emergency has emphasized the need to study SARS-CoV-2 pathogenesis. The human microbiome has been shown to regulate the host immune system and may influence host susceptibility to viral infection, as well as disease severity. Several studies have assessed whether compositional alterations in the nasopharyngeal microbiota are associated with SARS-CoV-2 infection. However, the results of these studies were varied, and many did not account for disease severity. This study aims to examine whether compositional differences in the nasopharyngeal microbiota are associated with SARS-CoV-2 infection status and disease severity.

**Methods:** We performed Nanopore full-length 16S rRNA sequencing on 194 nasopharyngeal swab specimens from hospitalized and community-dwelling SARS-CoV-2-infected and uninfected individuals. Sequence data analysis was performed using the BugSeq 16S analysis pipeline.

**Results:** We found significant beta (PERMANOVA p < 0.05), but not alpha (Kruskal-Wallis p > 0.05) diversity differences in the nasopharyngeal microbiota among our study groups. We identified several differentially abundant taxa associated with SARS-CoV-2 infection status and disease severity using ALDEx2. Finally, we observed a trend towards higher abundance of Enterobacteriaceae in specimens from hospitalized SARS-CoV-2-infected patients.

**Conclusions:** This study identified several alterations in the nasopharyngeal microbiome associated with SARS-CoV-2 infection status and disease severity. Understanding the role of the microbiome in infection susceptibility and severity may open new avenues of research for disease prevention and treatment.

## Introduction

The global COVID-19 pandemic and ensuing public health emergency has resulted in enormous economic costs and healthcare burdens worldwide. Over 500 million people have been infected and over 6 million people have died from COVID-19 (https://ourworldindata.org/covid-cases). COVID-19 is caused by severe acute respiratory syndrome coronavirus 2 (SARS-CoV-2); a positive-sense RNA virus of the *Coronaviridae* family [1]. There is wide variation in individual risk of SARS-CoV-2 infection and clinical outcomes; however, the mechanisms of SARS-CoV-2 pathogenesis and differences in COVID-19 disease progression remain unclear.

Associations between the human microbiome and the development of disease have been widely studied in recent years. The influence of the microbiome on viral infection and respiratory health has been explored [2], and the immunomodulatory role of the mucosal microbiome has been posed as a mechanism that may influence host susceptibility to viral infection [3,4]. Indeed, the presence of certain commensals have been shown to influence host Toll-like receptor expression, which is involved in virus detection and immunity [5]. Elevated nasal and systemic levels of pro-inflammatory cytokines have also been implicated in adverse clinical outcomes in influenza patients [5]. Therefore, microbes that overstimulate host cytokine responses may influence viral infection severity.

Active viral infection has also been shown to alter the composition of the respiratory microbiota [6] and influenza infection may increase host susceptibility to bacterial co-infections [7]. Several studies have assessed the prevalence of bacterial co-infections in SARS-CoV-2 patients, with one systematic review estimating the prevalence of co-infections in COVID-19 patients at 6.9% [8]. As bacterial co-infections can lead to adverse patient outcomes, it is important to investigate whether there are associations between SARS-CoV-2 infection and disease severity, and the composition of the upper respiratory tract microbiome.

Several studies have examined the relationship between SARS-CoV-2 infection and the oral, nasal, lung, and gut microbiomes. However, the results of these studies have varied greatly. *Pseudomonas aeruginosa* [9], *Fusobacterium periodonticum* [10], and Propionibacteraceae [11] were all found to be differentially abundant in samples from SARS-CoV-2-infected patients versus healthy controls. Additionally, several studies have shown decreased alpha diversity in the nasal microbiome in SARS-CoV-2-infected patients compared to healthy controls [11,12]. However, other groups reported no decrease in alpha diversity metrics for SARS-CoV-2-infected patients [9,10], and one study found an increase in species richness among samples from SARS-CoV-2-infected patients [13]. These studies differ in their sample handling, subject inclusion criteria, and study groups definitions, which makes direct comparisons between studies difficult. Furthermore, many of these studies had limited small sample sizes and did not account for disease severity, viral load, collection date, or SARS-CoV-2 variant type, which may result in confounded study results. The relationship between SARS-CoV-2 infection and the nasopharyngeal microbiome composition remains unclear.

In this study, we will harness full-length 16S rRNA sequence data to assess compositional differences in the nasopharyngeal microbiota between four groups of patients: SARS-CoV-2-infected-hospitalized patients, SARS-CoV-2-infected community-dwelling patients, SARS-CoV-2-uninfected-hospitalized patients, and SARS-CoV-2-uninfected community-dwelling patients from British Columbia.

## Materials & Methods

### Study population and specimen collection

Nasopharyngeal swab (NPS) specimens (n = 194) from adult individuals, collected in Copan Universal Transport Medium (Copan, Murrieta, CA) or Yocon Viral Transport Medium (Yocon Biology, Beijing, China), were tested for SARS-CoV-2 at the Vancouver General Hospital (VGH) Division of Medical Microbiology. Study samples were collected retrospectively throughout the COVID-19 pandemic in British Columbia (March 2020 – January 2022) and distributed across four study groups, SARS-CoV-2-uninfected community-dwelling (COMNEG) (n = 51), SARS-CoV-2-infected community-dwelling (COMPOS) (n = 47), SARS-CoV-2-uninfected hospitalized (HOSNEG) (n = 48), and SARS-CoV-2-infected hospitalized (HOSPOS) (n = 48) specimens.

Routine diagnostic testing was performed using either a Roche MagNA Pure extraction system (Roche Diagnostics, Laval, Canada) in combination with a lab developed (LDT) real time PCR (RT-PCR) assay detecting the E-gene and RdRp gene targets, or the Panther Fusion SARS-CoV-2 assay (Hologic Inc., San Diego, CA), detecting two targets in ORF1ab. The LDT RT-PCR assay was developed at the British Columbia Center for Disease Control (BCCDC) and is based on the World Health Organization’s guidelines for COVID-19 RT-PCR diagnostic screening [14]. RT-PCR C_t_ values were available for most SARS-CoV-2 positive specimens. Several NPS underwent testing using the BioFire respiratory panel 2.1 (Biomerieux, St-Laurent, Canada), C_t_ values for these specimens were not available. Screening for potential SARS-CoV-2 variants of concern (VOCs) (e.g. alpha, beta, gamma, delta, omicron) was performed using PCR at the BCCDC using an assay developed in-house [no reference or report (or web URL) for this].

### Nucleic Acid Extraction & Quantification

Total nucleic acids were extracted from NPS specimens using the MagNA Pure 24 Total NA Isolation Kit on the MagNA Pure 24 extraction system (Roche Diagnostics, Laval, Canada). The Pathogen 1000 protocol was used with a 500µL sample and 50µL elution volume. Total nucleic acid extracts were quantified with a Qubit 4 Fluorometer (Thermo Fisher Scientific) using the high-sensitivity dsDNA assay kit.

### Library Preparation & Sequencing

We utilized the Oxford Nanopore Technologies (ONT) MinION sequencing platform for this study, which enables real-time, long read sequencing of biological samples. This approach may confer more accurate taxonomic resolution than short-read 16S rRNA sequencing approaches [15]. Library preparation was performed using the full-length 16S rRNA barcoding kit (ONT: SQK-16S024), with several key modifications. Briefly, 20ng of nucleic acid extract (up to 10µL) was combined with barcoded primers (ONT), LongAmp HotStart Taq 2x Master Mix (New England Biolabs), and PCR-grade water as specified in the SQK-16S024 protocol (ONT). Full-length 16S amplicon fragments were amplified through PCR, followed by magnetic bead cleanup using 30µL of PCRClean DX beads (Aline Biosciences). Amplified libraries were quantified as described above and 2ng of each sample (up to 2µL) was pooled together. Up to 23 clinical samples plus a negative control (extracted blank viral transport medium) were multiplexed per flowcell. Adaptor ligation was performed at room temperature using 15µL of pooled library, followed by addition of SQB and Loading Beads (ONT). Final libraries were loaded onto MinION flowcells (FLO-MIN106) and sequenced using default run parameters in MinKNOW (Version 4.2.8, ONT) for up to 72 hours.

### Sequence Data Analysis

Raw sequence data were basecalled using Guppy (Version 5.0.7, ONT) with default parameters and the – *device cuda:0* flag to enable GPU basecalling. Basecalled fastq files were analyzed using the BugSeq 16S sequencing analysis pipeline [16]. Downstream analysis and visualization of Amplicon Sequence Variant (ASV) classification tables was performed using Rstudio (R Version 4.1.0) [17]. Relative abundance was calculated for each taxon and alpha/beta diversity metrics were estimated using the *vegan* R package [18]. Multivariate analyses (Principal coordinates analysis (PcoA), and permutational multivariate analysis of variance (PERMANOVA)) were performed to assess compositional differences in the nasopharyngeal microbiome among our four study groups. ALDEx2 [19] was used to contrast differential abundance of taxa across our study groups.

### Ethics Approval

This study included de-identified samples collected from VGH and has obtained ethics board approval from the research ethics board at the University of British Columbia (H20-02152).

## Results

### Study Population

Our study groups exhibited similar age distributions (Table 1); however, there were a larger number of male subjects included in the study (39.7% female, 60.3% male). Study samples were collected from March 2020 to January 2022, providing access to clinical specimens positive for a range of SARS-CoV-2 lineages and representing all seasons (Table 1; Supplementary Figure 1).

**Table 1.** Summary of study population demographics.

|  | Male (n) | Female (n) | Age | Patient location | Specimen collection |
| --- | --- | --- | --- | --- | --- |
| COMNEG | 24 | 27 | 60.9 ± 12.2 | Community-dwelling | Mar-May 2020, n = 1<br>Aug-Dec 2020, n = 3<br>Jan-May 2021, n = 29<br>Aug 2021-Jan 2022, n = 18 |
| COMPOS | 27 | 20 | 61.7 ± 13.6 | Community-dwelling | Mar-May 2020, n = 0<br>Aug-Dec 2020, n = 23<br>Jan-May 2021, n = 23<br>Aug 2021-Jan 2022, n = 1 |
| HOSNEG | 33 | 15 | 61.9 ± 14.7 | Non-critical care unit, n = 44<br>Critical care unit, n = 4 | Mar-May 2020, n = 0<br>Aug-Dec 2020, n = 0<br>Jan-May 2021, n = 43<br>Aug 2021-Jan 2022, n = 5 |
| HOSPOS | 33 | 15 | 64.1 ± 15.1 | Non-critical care unit, n = 12<br>Critical care unit, n = 36 | Mar-May 2020, n = 11<br>Aug-Dec 2020, n = 10<br>Jan-May 2021, n = 26<br>Aug 2021-Jan 2022, n = 1 |

**Fig 1.**
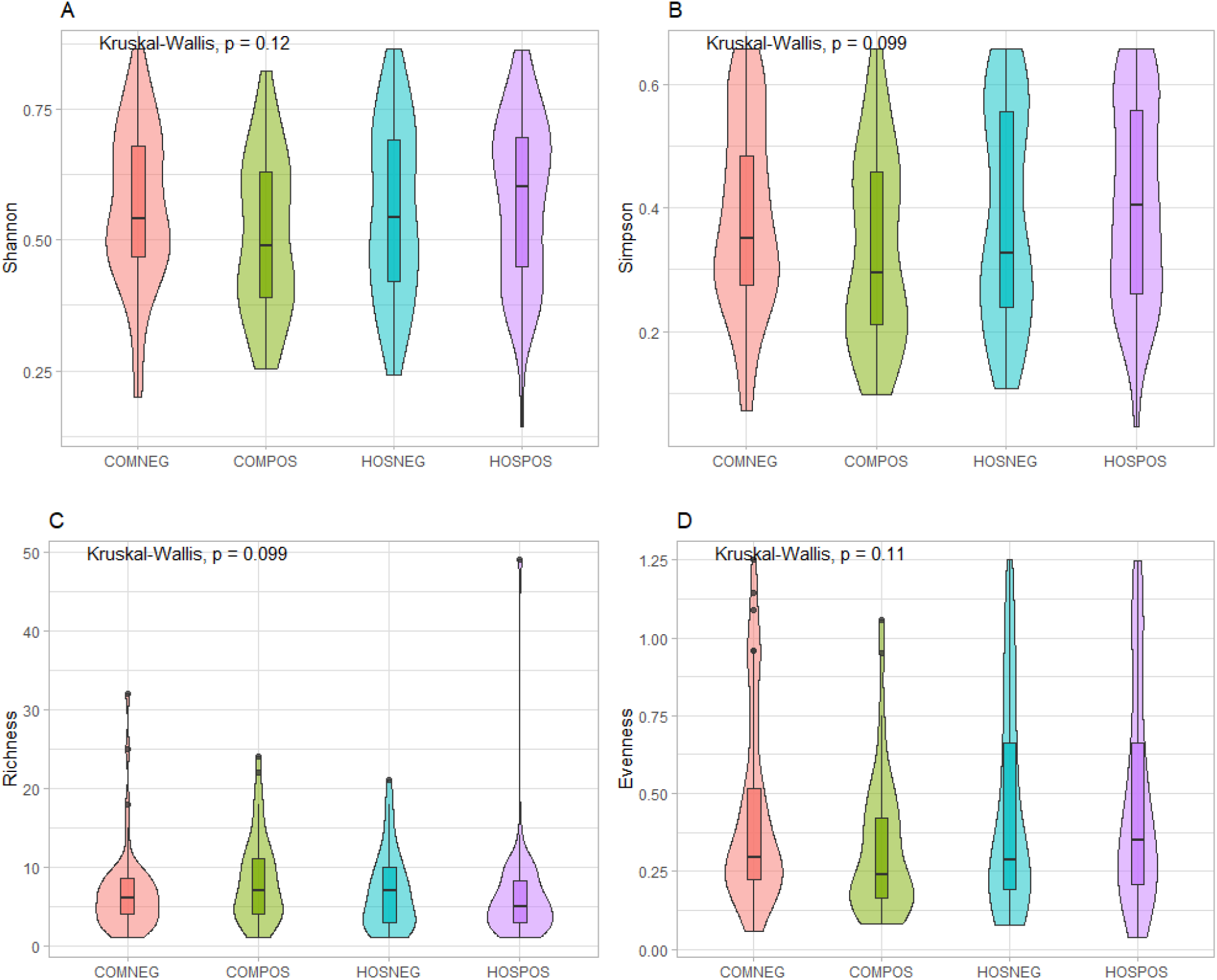
Genus-level differences in alpha diversity among our study groups summarized through **A**. Shannon diversity index, **B**. Simpson diversity index, **C**. Taxa richness, **D**. Evenness.

### Richness, Evenness, & Diversity Metrics

We did not observe any significant differences in alpha diversity among our study groups (Kruskal-Wallis p > 0.05) (Fig 1), although there did appear to be a trend towards lower mean Shannon/Simpson diversity and evenness for SARS-CoV-2-infected community-dwelling samples versus SARS-CoV-2-uninfected community-dwelling samples. This trend was not observed in hospitalized SARS-CoV-2-infected versus uninfected samples.

Bray-Curtis dissimilarity ordination was performed to assess differences in beta diversity among our study groups using PCoA at the species-, genus-, and family-level taxonomic ranks (Fig 2). Statistical analysis was performed to determine whether there was significant clustering among our study groups using *adonis* within the *vegan* R package. We observed significant clustering at all three taxonomic ranks (adonis: Species p = 0.009, Genus p = 0.028, Family p = 0.027), suggesting that there are differences in microbial community composition among our study groups, although, there was a large degree of overlap in the 95% confidence ellipses.

**Fig 2.**
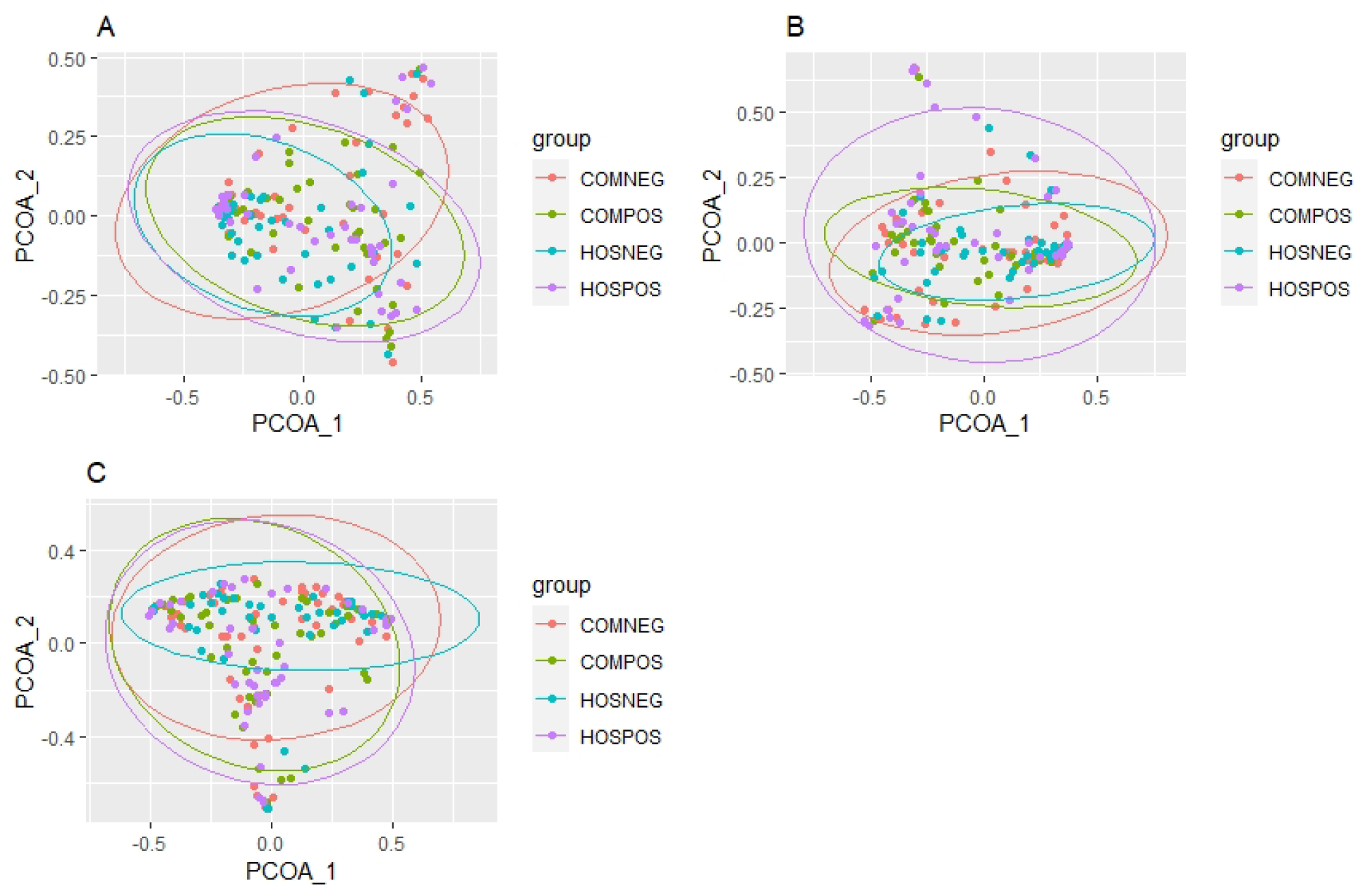
Principle coordinates analysis plots based on Bray-Curtis dissimilarity among our four study groups at the **A**. species, **B**. genus, and **C**. family taxonomic ranks. P-values were derived from PERMANOVA.

### Taxonomic Differences Among our Study Groups

Relative abundance was calculated, and taxa present at less than 5% prevalence across all samples were removed. At the genus level, *Staphylococcus* was the most abundant genus on average for both SARS-CoV-2-uninfected study groups, whereas *Acinetobacter* was the most abundant genus in the hospitalized infected group and *Moraxella* was the most abundant genus in the community-dwelling SARS-CoV-2-infected group (Supplemental Figure 2). At the species level, both SARS-CoV-2-infected groups were dominated by common nasal pathobionts and opportunistic pathogens including *Haemophilus influenzae, Staphylococcus haemolyticus*, and *Staphylococcus aureus*, with the SARS-CoV-2-infected hospitalized group having the opportunistic pathogen *Klebsiella aerogenes* as the most abundant species (Fig 3). At the family rank, SARS-CoV-2-infected hospitalized patients displayed a higher mean relative abundance and broader range of Enterobacteriaceae then any of the other study groups (Supplemental Figure 3).

**Fig 3.**
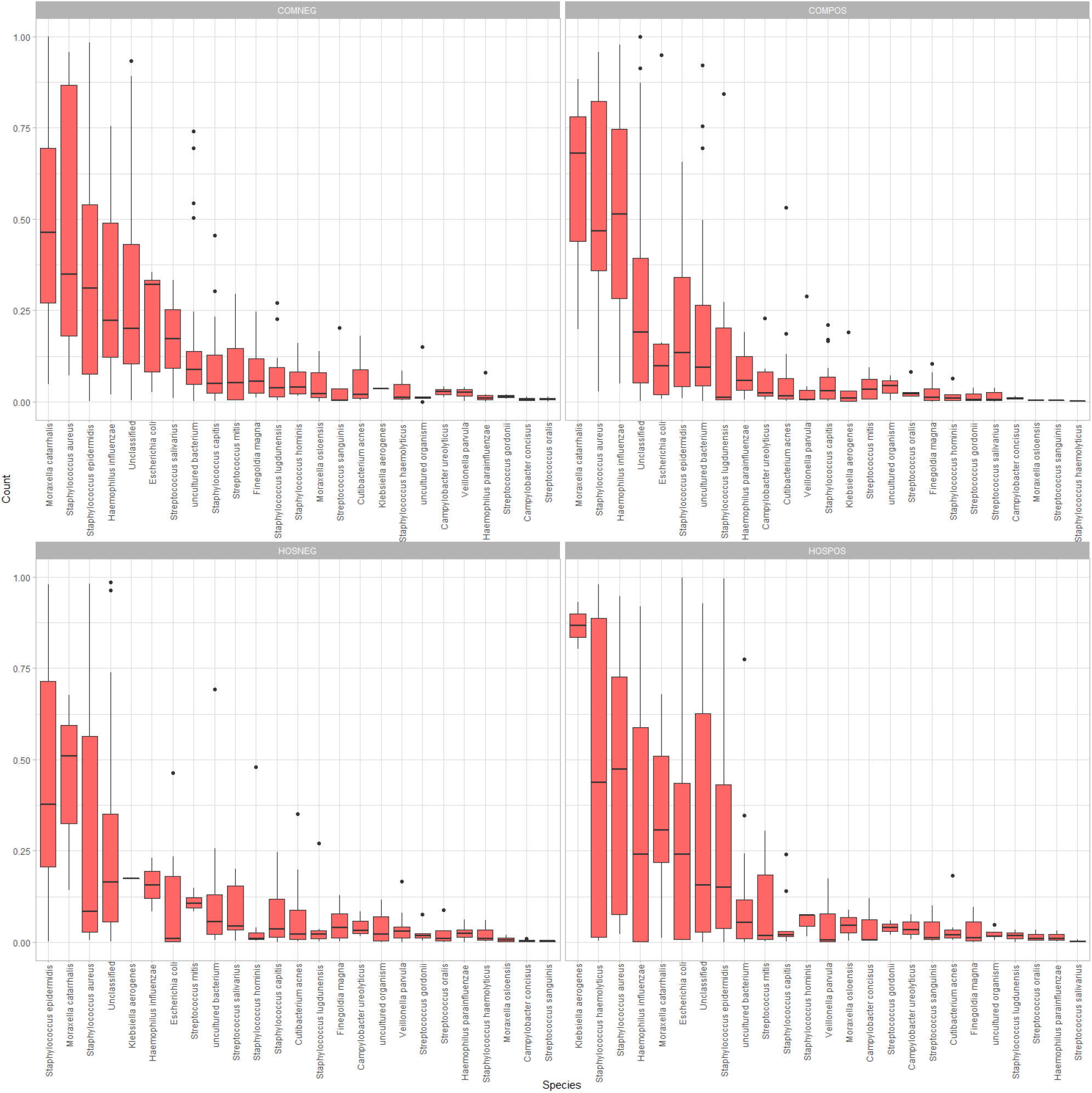
Side-by-side boxplots of relative abundance at the species-level among our four study groups.

ADLEx2 was used to assess which of the taxa were significantly differentially abundant. The species *Cutibacterium acnes*, the genera *Cutibacterium* and *Peptinophilus*, and the families Propionibacteriaceae and Peptostreptococcales-Tissierellales were all differentially abundant over the four study groups (Kruskal-Wallis p < 0.05). An “uncultured” genus assigned to the family Neisseriaceae was also found to be differentially abundant. All the differentially abundant taxa were enriched in community-dwelling SARS-CoV-2 infected individuals.

### Alterations in the Nasopharyngeal Microbiome Associated with C_t_ Value, Collection Date, and Variant Status

Significant beta (PERMANOVA p < 0.05), but not alpha diversity differences in the nasopharyngeal microbiome composition were observed when stratified by sample collection date (Fig 4). ALDEx2 revealed significant differences (Kruskal-Wallis p < 0.05) in *Streptococcus spp*. and *Corynebacterium spp*. among samples from infected individuals collected during the first three waves of the pandemic in British Columbia (Wave 1: March-May 2020, Wave 2: August-December 2020, Wave 3: January-May 2021). We did not observe significant differences in alpha (Kruskal-Wallis p > 0.05), or beta diversity (*adonis* p > 0.05) associated with viral load or by SARS-CoV-2 variant infection status.

**Fig 4.**
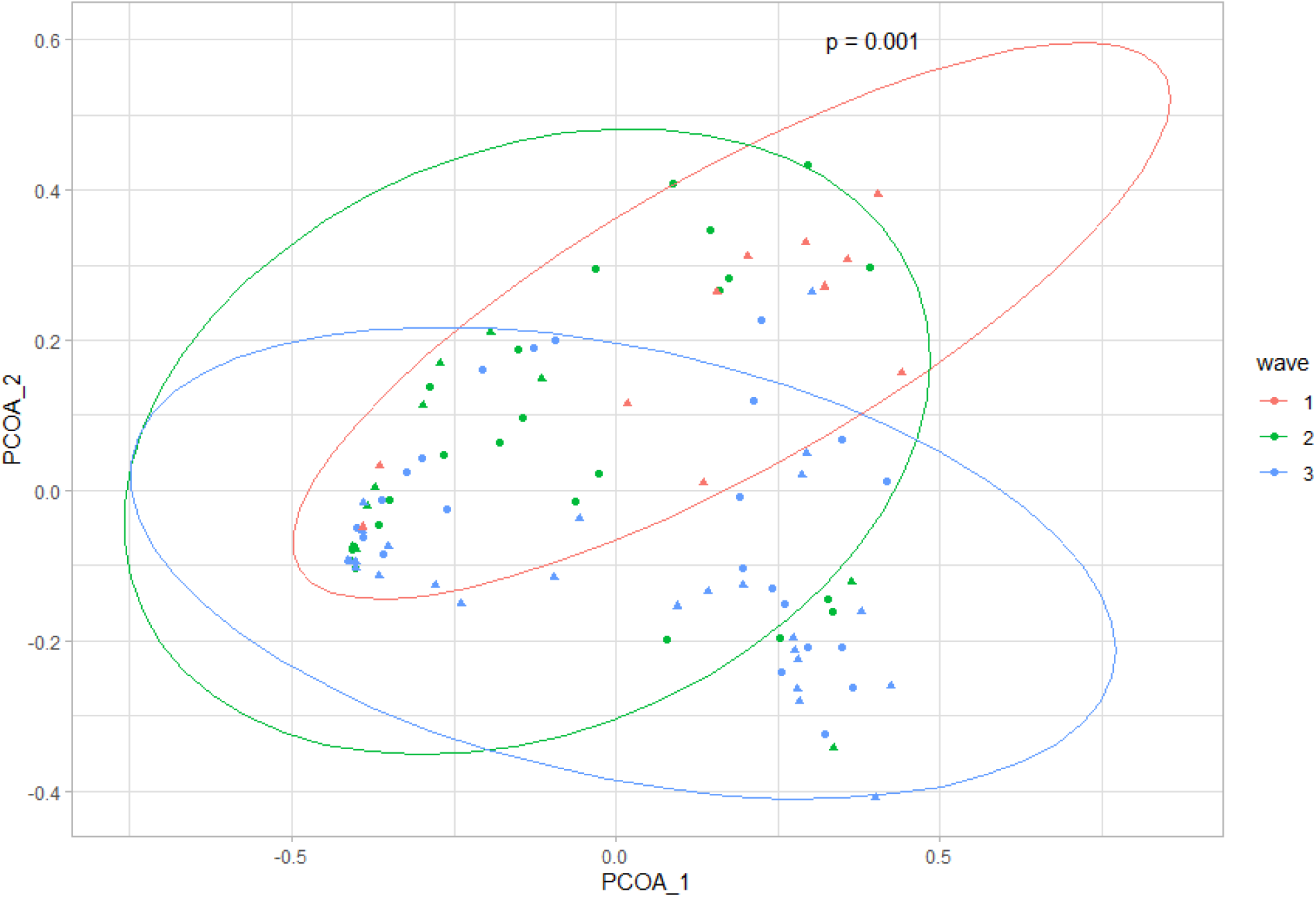
PCoA plot for genus-level differences in the nasopharyngeal microbiome based on Bray-Curtis dissimilarity stratified by collection date. Circles indicate community-dwelling SARS-CoV-2-infected subjects and triangles indicate hospitalized SARS-CoV-2-infected subjects.

## Discussion

In this study, we used full-length 16S rRNA sequencing data to examine differences in the nasopharyngeal microbiota associated with SARS-CoV-2 infection and COVID-19 severity. We found significant beta, but not alpha diversity differences in the nasopharyngeal microbiota between hospitalized and community-dwelling SARS-CoV-2-infected and uninfected specimens. Our results are consistent with two previous studies that found no significant difference in alpha diversity between SARS-CoV-2-infected and uninfected patient samples [9,10]. Both studies displayed a similar trend towards decreasing alpha diversity in SARS-CoV-2 patients, consistent with our data. Our alpha diversity observations contrast with two previous studies [11,12] that found a decrease in alpha diversity among SARS-CoV-2 infected specimens; however, these studies did not account for disease severity. We found significant differences in beta diversity among our study groups, consistent with other groups that have found significant beta diversity clustering between SARS-CoV-2-infected and uninfected subjects [9,10,11]. However, we did see significant overlap in the confidence ellipses, suggesting that although significant, the magnitude of compositional differences in the microbiota among our study groups are relatively small.

We identified several trends in the taxonomic composition of the nasal microbiota among our study groups, as well as a few differentially abundant taxa. The high relative abundance of *Staphylococcus spp*. among our SARS-CoV-2-uninfected groups is consistent with literature describing the core nasal microbiota among adults that is typically dominated by the phyla Actinobacteria and Firmicutes, including *Staphylococcus* [20]. At the species level, the trend towards non-statistically significant higher mean abundance of opportunistic pathogens and pathobionts such as *Haemophilus influenzae, Staphylococcus haemolyticus, Staphylococcus aureus*, and *Klebsiella aerogenes* among our SARS-CoV-2-infected specimens, is consistent with literature implicating some of these opportunistic pathogens in respiratory virus infection severity [5]. In particular, the presence of *H. influenzae* in the nasopharynx has been shown to promote human rhinovirus pathogenesis through the increased expression of pulmonary epithelial cell TLR-3 [21]. Additionally, non-influenza respiratory viral infection has been hypothesized to increase host susceptibility to *S. aureus* superinfection through alterations in host-*S. aureus* adhesion, increased epithelial cell permeability to *S. aureus*, or a reduction in the immune system’s ability to regulate *S. aureus* clearance from the nasal passage [22], in line with the trend we saw towards higher *S. aureus* abundance in our SARS-CoV-2-infected study groups. The trend towards increased abundance of Enterobacteriaceae among our hospitalized SARS-CoV-2-infected study group may be attributed to medical interventions in this group such as, intubation or antibiotic exposure. We found differential abundance of *Cutibacterium acnes* among our study groups. This nasal commensal has been implicated in patients with chronic rhinosinusitis [23] and is associated with increased host inflammatory response [24], which may explain why this species was most abundant in our community SARS-CoV-2-infected group. This species has also been hypothesized to play a role in regulating nasal microbiome homeostasis through its’ ability to influence *S. aureus* growth [25]. Finally, the presence of an “uncultured” Neisseriaceae member that was most abundant in our community-dwelling SARS-CoV-2-infected group is interesting. This family has been previously reported as a “core” nasopharyngeal microbiome taxon [26] and has been shown to be associated with swine influenza co-infection [27]. However, it’s role in SARS-CoV-2 infection remains unclear.

Our study has several limitations. First, our samples were de-identified, therefore, we were unable to adjust for potential confounders that may have influenced the nasopharyngeal microbiota among our study groups such as medications, medical interventions, or hospitalization duration in our hospitalized study groups. SARS-CoV-2-uninfected community-dwelling individuals may have other respiratory viral infections. Without knowledge of detailed symptoms that led individuals to seek COVID testing, it is impossible to determine if or how many of our negative samples represent true viral infection negative samples. This may have led to an underestimate in the magnitude of our alpha and beta diversity estimate differences across groups. However, the incidence of other respiratory viruses was low in our patient population during the study period (https://www.canada.ca/en/public-health/services/diseases/flu-influenza/influenza-surveillance.html). We identified a high proportion of reads mapping to the genus *Alishewanella*, an environmental microbe. This may be due to poor annotation of this taxa in the reference database. Functional analysis from metatranscriptomic data examining this relationship may reveal functional pathways that are differentially expressed in SARS-CoV-2 infection. Microbiome function may play a more significant role than taxonomic differences in infection progression and severity.

The results of this study present several key alterations in the nasopharyngeal microbiota associated with SARS-CoV-2 infection and disease severity. These results emphasize the possible role of the respiratory microbiome in host susceptibility to viral infection and subsequent disease severity. Our study accounted for COVID-19 severity and harnessed full-length 16S rRNA sequence data that may provide a more granular representation of the microbiota differences in SARS-CoV-2-infected and uninfected individuals. Further work is necessary to determine whether functional characteristics of the nasopharyngeal microbiome are associated with respiratory viral infection and adverse patient outcomes, or if emerging SARS-CoV-2 variants have alternate influences on the nasal microbiota. Understanding these microbiome-driven mechanisms could present novel prognostic markers, or offer new approaches to disease prevention and treatment.

## Data Availability

All data produced in the present study are available upon reasonable request to the authors

