## Supplementary Tables/Figures for "Alterations in the Nasopharyngeal Microbiome Associated with SARS-CoV-2 Infection Status and Disease Severity"

**SUPPLEMENTARY MATERIAL**

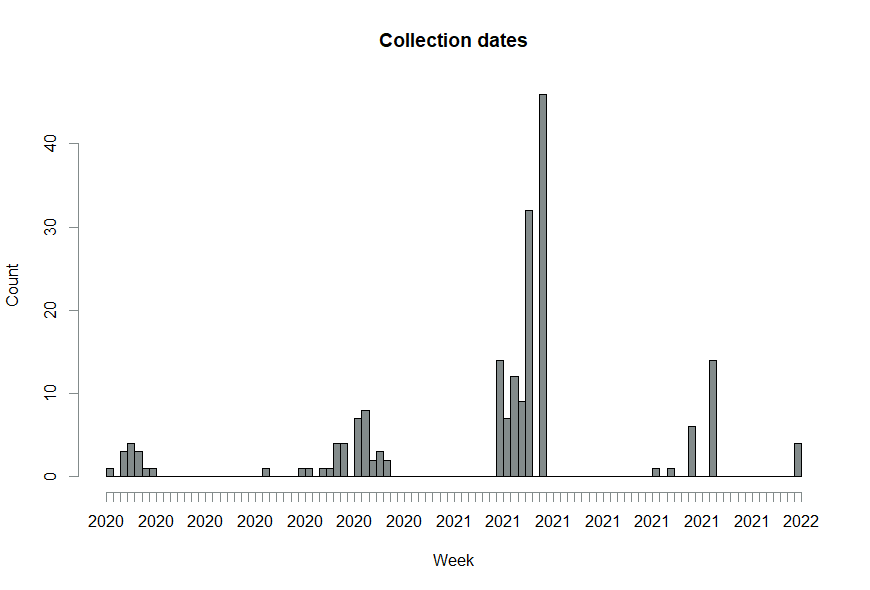

**S1.** Histogram of collection dates for study specimens.

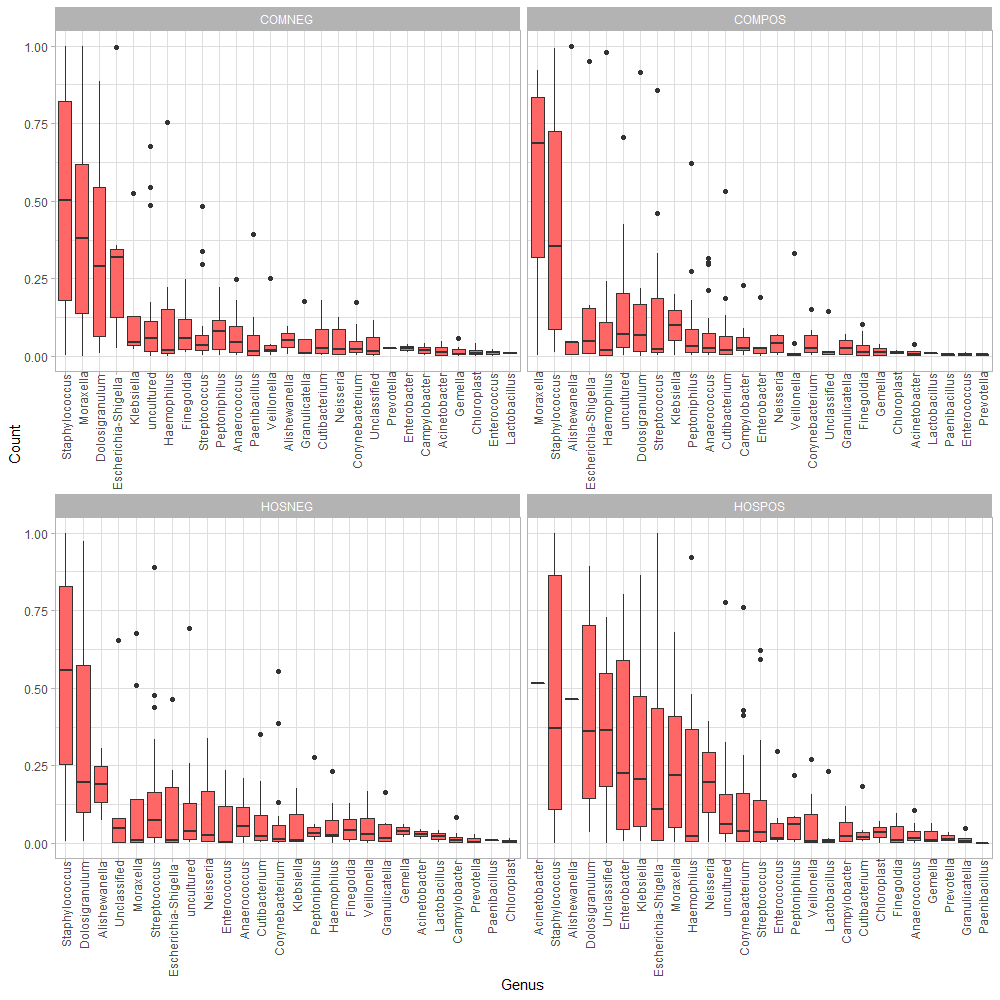

**S2.** Side-by-side boxplots of relative abundance at the genus-level among our four study groups.

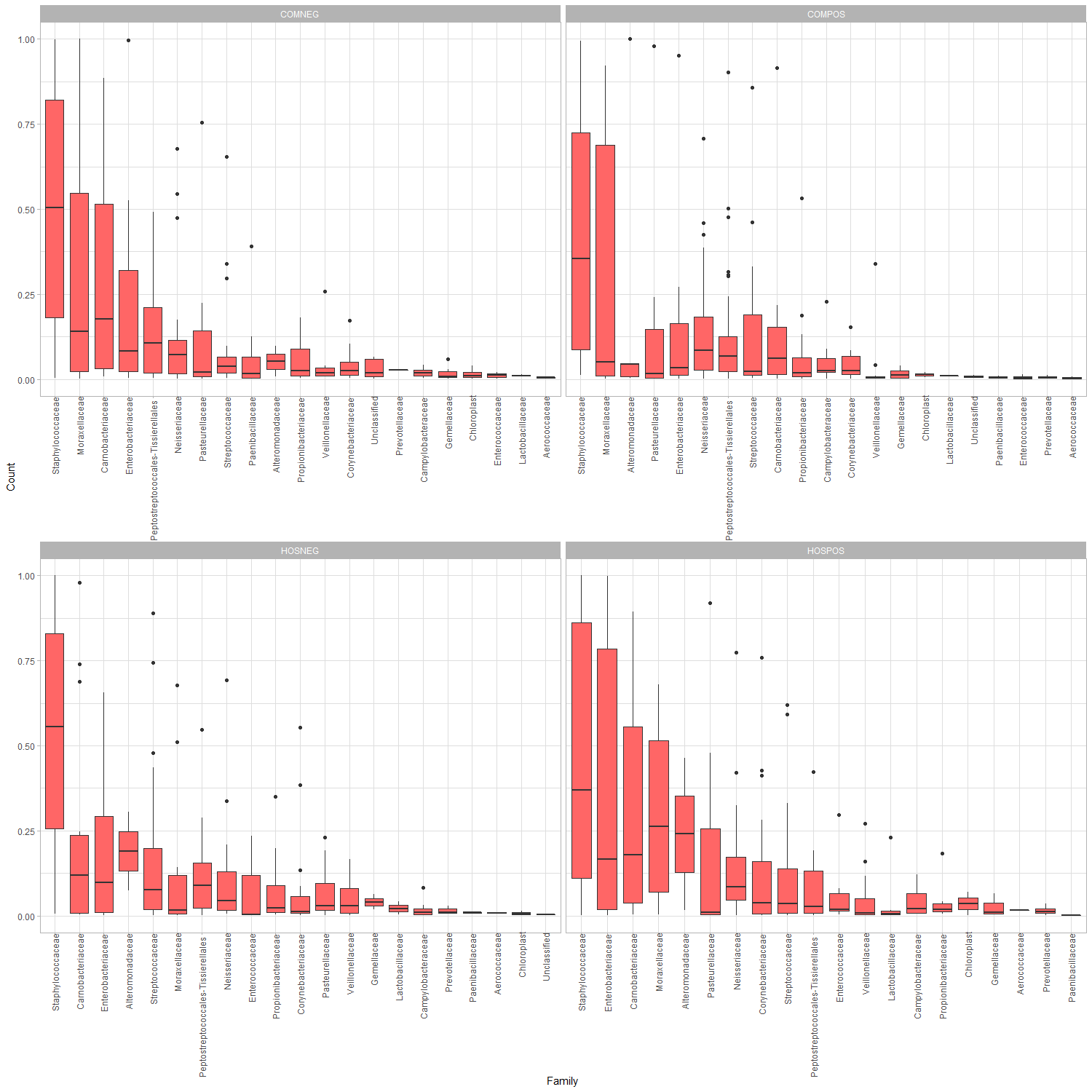

**S3.** Side-by-side boxplots of relative abundance at the family-level among our four study groups.
